# GAMBIT: A Digital Tool to Train Distinct Inhibitory Control Mechanisms

**DOI:** 10.64898/2026.03.05.26347639

**Authors:** G. Dirupo, M.L. Westwater, S. Khaikin, A. Feder, J.M. DePierro, D.S. Charney, J.W. Murrough, L.S. Morris

## Abstract

Deficits in inhibitory control are common across a wide range of psychiatric disorders and are closely linked to symptom severity, including emotional dysregulation, anxiety, substance misuse, and self-harm, making them an appealing target for intervention. Cognitive training offers a low-cost, scalable, and non-invasive strategy to strengthen inhibitory control; however, most existing paradigms target only a single facet of inhibition and rarely account for environmental influences, such as affective context. To address these gaps, we developed a computerized inhibitory control training paradigm to simultaneously engage three components of inhibition: preemptive, proactive, and reactive, while embedding trials within positive and negative affective contexts to assess the impact of emotional stimuli. Across two online experiments, participants completed the GAMBIT task in one session (Experiment 1, N = 300) or repeated over three sessions (Experiment 2, N = 65). The task included No-Go trials to train preemptive inhibition, stop-signal trials for reactive inhibition, and stop-signal anticipation trials to train proactive inhibition. Affective images of differing valence were presented as background stimuli to evaluate their impact on inhibitory performance. In Experiment 1, participants showed higher accuracy on No-Go versus reference Go trials (β=1.45, SE=0.09, p<.001), confirming successful manipulation of preemptive inhibition. Reaction times were slower during anticipation trials across two different conditions (β=0.16, SE=0.04, p<.001; β = 0.07, SE = 0.04, p = 0.047), consistent with proactive slowing when anticipating a potential stop signal. Additionally, positive affective images (β = 0.10, SE= 0.009, p < 0.001) further slowed RTs, indicating emotional interference with proactive control. In Experiment 2, the pattern of higher No-Go accuracy was replicated (β = 0.91, SE = 0.11, p < .001) and accuracy generally improved over sessions (β = 0.38, SE = 0.06, p < .001). In anticipation trials, RTs become shorter across sessions (session 2: β = -0.25, SE = 0.06, p < .001; session 3: β = -0.45, SE = 0.06, p < .001), reflecting practice-related gains, and SSRTs decreased over time (F_(2,56)_ = 6.26, p = .004), consistent with enhanced reactive inhibition. Proactive inhibition was modulated by affective images, with both negative (β = 0.04, SE = 0.02, p = .039) and positive (β = 0.16, SE = 0.02, p < .001) affective images associated with slower RTs. Participants also reported reductions in self-assessed temper control by the last session (W = 25.5, p = .007, q = .037, d = -0.51) and usability ratings were high (all means ≥ 3.87/5). Together, these findings show that this paradigm recruits multiple forms of inhibitory control and yields training-related improvements in both performance and affective outcomes. This provides preliminary validation of a scalable, fully online inhibitory control training tool targeting multiple dissociable inhibitory processes within affective contexts. The approach holds promise as an accessible transdiagnostic intervention to support symptom improvement across psychiatric disorders, with future work needed to evaluate clinical efficacy in patient populations.

## 1. Introduction

Inhibitory control deficits are present across multiple psychiatric disorders ^1^, and are associated with poorer functional outcomes across psychopathologies ^2,3^ and greater severity in a range of symptoms. Those include impaired regulation of emotional responses, difficulty suppressing intrusive thoughts or memories ^4^, and heightened impulsivity ^5^, making inhibitory control an appealing therapeutic target across psychiatric conditions. Although inhibitory control is defined as the inability to stop, change, or delay behaviors that are not appropriate for the context ^5^, it is a multi-faceted construct that includes different components such as: preemptive inhibition, the ability to stop immediately; reactive inhibition, the ability to stop after a delay; and proactive inhibition, the ability to anticipate the need to stop based on probabilistic context cues ^6^. This is also supported by biological evidence, with a recent large-scale genetic study finding that distinct impulsivity facets show unique genetic architecture ^7^, neural evidence reporting distinct cortico-striatal substrates for impulsivity related to the temporal discounting of reward, motor or response disinhibition ^8^ and overall micro and macro neural cross species mechanisms ^9^.

Importantly, better behavioral control is associated with improvement in psychiatric symptoms. For example, psychosocial interventions targeting cognitive and behavioral regulation, including cognitive-behavioral therapy (CBT) and related approaches, have been shown to reduce self-harm frequency and severity while also alleviating depression and anxiety, and improving self-regulation ^10,11^. However, CBT and other psychotherapeutic approaches can be costly, time-intensive, and dependent on specialized expertise, thus limiting accessibility, particularly for individuals in rural areas ^12^. Scalable interventions are therefore needed to reach individuals traditionally underrepresented in clinical settings and lacking access to mental health services ^13^. Digital psychiatric interventions hold considerable promise for expanding access to care ^14,15^. Evidence from self-management interventions suggests that individuals with elevated depression or anxiety symptoms may show especially strong gains in empowerment and symptom-related outcomes when behavioral control strategies are engaged ^16^. Together, these findings support a close link between improvements in behavioral control processes and meaningful changes in clinical symptoms.

Being a low-cost and non-invasive technique for effectively modifying behavioral biases, digital cognitive training is increasingly used in clinical and research settings ^14^. Prior work has shown that cognitive training tasks grounded in neuropsychological models of psychiatric disorders hold promise as novel interventions. For example, the emotional faces memory task (EFMT), now marketed as *ReJoyn*, is the first FDA-approved digital therapeutic for depression ^17^, with proven changes in symptoms as well as brain functions associated with its use and symptom changes ^18^. As described above, inhibitory control has emerged as a promising therapeutic target due to its responsiveness to interventions across psychiatric conditions ^19^. For instance, adaptive Go/No-Go training is associated with improved response inhibition and symptom alleviation in students with PTSD symptoms ^20^. Despite these findings, additional aspects of training effectiveness and its usability remain to be addressed. For example, critically, current interventions largely conceptualize inhibitory control as a unitary mechanism, neglecting to target its dissociable cognitive and affective components. Moreover, while it is known that the valence of affective context differentially impacts performance in healthy individuals ^21^ and, to a greater extent, in psychiatric populations ^22^, this element is rarely incorporated into training paradigms.

Here, we introduce a novel computerized task designed to capture and train three distinct components of inhibitory control: preemptive, reactive, and proactive. We provide evidence that validates our novel, fully computerized, gamified cognitive training tool: Gamified Approach to Maximizing Biobehavioral Inhibition in Transdiagnostic inhibition-related disorders (GAMBIT). By adopting a transdiagnostic framework, this approach aims to address a core biobehavioral processes implicated across a broad range of psychiatric conditions rather than disorder-specific manifestations. We aimed to investigate whether GAMBIT can successfully capture different aspects of inhibitory control (Experiment 1) and whether repeated engagement with GAMBIT is associated with (i) better task performance (ii) reduced inhibitory control related psychiatric symptoms (Experiment 2). The GAMBIT task is low-cost, scalable, and deliverable entirely online, thereby directly addressing key barriers to treatment accessibility, including cost, availability, and participant or clinician burden. Our tool has the potential to complement existing interventions and extend access to evidence-based cognitive training. As the present study focuses on task validation in a non-clinical sample, future work will be required to evaluate its clinical efficacy and symptom-level impact in patient populations.

## 2. Methods

To test our hypothesis, we conducted two independent, fully online experiments. In Experiment 1, a sample of n=302 participants completed one single session. In Experiment 2, n=65 participants completed the same task across three separate sessions within one week.

### 2.1 Procedure

All participants were recruited remotely through Prolific (**Error! Hyperlink reference not valid.** a platform providing a verified and high-quality pool of research subjects. After recruitment, for both Experiment 1 and 2, participants were redirected to the Research Electronic Data Capture (REDCap) platform, where they completed a serie of self-report questionnaires (see Section 2.2.1 for full list). Upon completion, they were automatically redirected to the Pavlovia platform, which supports the online administration of cognitive tasks built with PsychoPy (v2024.2.4). Finally, after completing the task, they were redirected back to Prolific to receive a completion code finalizing their participation. Only for Experiment 2, after completing the first session, participants were instructed to return to Prolific independently for the second and third session. Each participant was enrolled in Experiment 2 on a Monday, and sessions 2 and 3 were available starting from Wednesday and Friday of the same week. After their last session, participants were asked to return to Prolific to complete post-task questionnaires on user experience. Longitudinal data collection was implemented using Prolific’s “Allowlist” feature, which enables the invitation of specific participants via their anonymized Prolific ID. Throughout this process, each participant’s unique Prolific ID was securely transmitted via embedded URL links, ensuring continuity across platforms while preserving anonymity. Participants received compensation of $10.50 per hour for each session across both experiments. The experiments were approved by the local ethics committee (Institutional Review Board approval: 21-00996).

### 2.2 Materials

#### 2.2.1 Self-reported measures

Before starting the task, participants completed a series of self-report questionnaires. These included sociodemographic information (e.g., age, sex, gender, education), global health ratings, and selected items from the Patient-Reported Outcomes Measurement Information System (PROMIS) questionnaire assessing multiple dimension (primary outcomes: anxiety and temper control; exploratory outcomes: alcohol use, positive affect, depression, and cognition) over the past week. Responses were recorded on a 4-point Likert scale (1 = “Never”, 4 = “Often”). In Experiment 2, participants completed the same scales at the end of session 3 as well as the PANAS questionnaire both before Session 1 and after Session 3. Finally, they completed a usability questionnaire at the end of the last session that consisted of four items measuring task difficulty, ease of understanding instructions, level of frustration, and willingness to participate again, which were answered on a 5-point Likert scale (1 = “Very slightly” or “Not at all”, 4 = “Extremely”). Finally, across both experiments, each session included one attention check question (e.g. “To confirm that you are paying attention, please select answer 3”).

#### 2.2.2 Task

GAMBIT is a novel computerized paradigm designed to simultaneously train different domains of inhibitory control. Each session included 520 trials, the majority of which were **Go trials** (n = 400), in which participants were instructed to respond as quickly as possible to a left- or right-positioned fish stimulus by pressing the “A” or “L” key on their keyboard. The other trial types manipulated three domains of inhibitory control: inhibition of response initiation (termed preemptive inhibition, No-Go trials), inhibition of an ongoing response (reactive inhibition, Stop-Signal trials), and anticipatory slowing based on contextual cues (proactive inhibition, Stop-Signal Anticipation trials) with high or low probability of a stop-signal. On **No-Go trials** (n = 40), a shark appeared in the center of the screen instead of the fish and participants were instructed to withhold their response. On **Stop-Signal trials** (n = 40), a tone was played for 1 second shortly after the fish appeared, signaling participants to withhold their response. An important metric in this task is the stop-signal delay (SSD), which refers to the individual time interval between the onset of the target stimulus (fish) and the stop signal (tone). This was dynamically adjusted throughout the task using a staircase model procedure to adapt the task difficulty to the individual’s performance. Specifically, SSD increased by 0.05 s following a successful stop and decreased by 0.05 s after an unsuccessful stop, starting at 0.05 s with a minimum threshold of 0.05 s and maximum of 0.8 s. This adaptive tracking approach was designed to maintain an overall stopping accuracy of approximately 50%, compensating for individual differences in response speed and inhibitory control. During **Stop-Signal Anticipation trials** (n = 80), the background color of the scene was a lighter or darker shade of green (as opposed to the light blue of all other trial types), indicating respectively a 25% (**low**, n=40) or 50% (**high**, n=40) probability that a stop signal (tone) would occur at a fixed delay of 0.2 s. These trials assessed proactive inhibition, by requiring participants to modulate their response based on a contextual cue (color of the background) predicting the likelihood of having to withhold their response. Most trials included emotionally valenced background images, equally including positive, negative and neutral affective context that were positioned in the corners of the screen and evenly distributed across trial types (see Fig. 1). Participants were instructed to respond as fast and accurately as possible across all trial types while ignoring the background images. Correct responses were rewarded with positive visual and auditory feedback (a gold coin animation). Following the task instructions, participants completed a comprehension quiz, and they were required to answer all questions correctly before proceeding to the task practice, otherwise they repeated the task instructions and quiz. Each session was preceded by a 40-trial practice block, consisting of only Go trials. Each full session lasted approximately 10 minutes.

**Figure 1.**
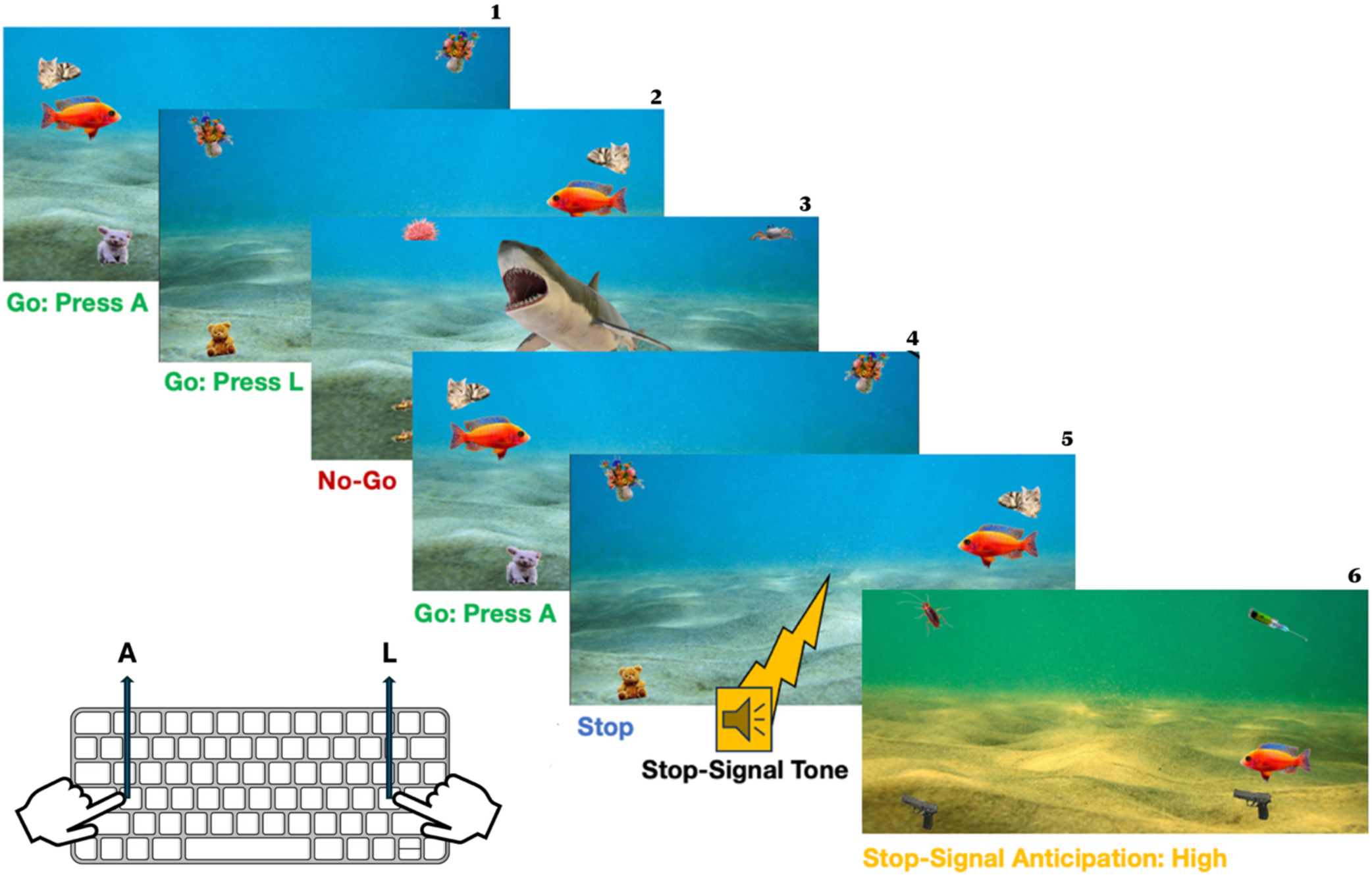
Schematic representation of the GAMBIT task. The task included four types of trials: Go trials (Example frames 1, 2, 4): a fish appeared on the left or right side of the screen, participants were instructed to respond with the corresponding keyboard press (“A” or “L”). No-Go trials (Example frame 3): a shark appeared instead of a fish, and participants were instructed to withhold their response. Stop trials (Example frame 5): a tone was presented after a variable delay following the fish stimulus, signaling participants to inhibit their response. Stop-Signal Anticipation trials (Example frame 6): the background color changed to indicate a 25% or 50% probability of a stop signal tone. Emotionally valenced images (positive, negative, neutral) were randomly assigned across all trials and visible in each example frame, at the corners of the screen.

### 2.3 Data Analyses

Analyses were performed using Python 3.12.4 (Anaconda distribution) and R 4.4.0 (model run via the libraries nlme and lme4) ^23^.

#### 2.3.1 Participant selection

Given the lack of live instructions and supervision, several selection filters were applied to ensure inclusion of only dedicated participants. First, participants who completed fewer than 400 trials were excluded (76.9% of the task). Then, those who provided incorrect answers to the attention checks (one question for Experiment 1, one question per session for Experiment 2 (total n=3), where in case of mistake we excluded only that session). Lastly, we excluded those who responded on fewer than 70% of Go trials (missed click, regardless of accuracy) and those with Stop trial accuracy <25% or >75% per guidelines in the field to obtain reliable SSRT metrics ^24^.

#### 2.3.2 Task Metrics

The primary task metrics analyzed were <u>accuracy</u> and <u>response time</u> (RT). Accuracy was defined based on whether a response (keyboard press) occurred when expected, and in case of Go trials, whether the correct key was pressed (“A” or “L”, corresponding to the fish location). For anticipation trials, when a keypress was expected, any response was considered correct, regardless of which key was pressed. RT was measured in milliseconds, calculated from stimulus onset to the participant’s keypress. Additionally, for each participant we computed the stop-signal reaction time (SSRT), i.e. the latency of the stopping process. SSRT was calculated subtracting the final SSD from the mean reaction time on Go trials and lower SSRT values indicate better reactive inhibition.

#### 2.3.3 Data preparation

Before performing the analyses, trials with response times (RTs) falling below the 25^th^ percentile and above the 75^th^ percentile of the RT distribution were removed to mitigate the influence of extreme values ^25,26^. Following outlier removal, RTs were transformed via the inverse rank method to reduce skewness and improve the distributional properties of the data ^27^ and in line with a recent report of a similar paradigm investigating inhibitory control ^28^. The outlier removal procedure was applied separately for each trial type and session to account for potential differences in distributions across conditions while the inverse rank transformation was applied without trial type distinction. Age in years was z-scored prior to analysis.

#### 2.3.4 Analyses

To examine differences in preemptive inhibition, we modeled accuracy at Go/ No-Go trials. Generalized linear mixed-effect models with a binomial distribution were used to compare Go and No-Go trial performance. Trial type was included as a fixed effect, with age and sex as covariates of no interest, and random intercepts for trial type were nested within the random effect of the participant. A response was considered correct for a No-Go trial when no keypress was detected, while for Go trials, there needed to be a keypress and the correct key needed to be clicked (A/ L depending on where the fish appeared on the screen). Accuracy in association to trial type including low and high anticipation trials and Stop trials as reference was also investigated (see Supplementary materials). In this case, a response was considered correct for high and low anticipation trials when a key was pressed (either A or L) when a response was expected (i.e. no stop sound was presented). Stop trials were correct when no keypress was detected.

To investigate proactive inhibition, differences in response times were estimated via a linear mixed-effects model using RTs as dependent variable. We included a main effect of trial type, and age and sex as covariates of no interest. The trial type term was a 3-level factor, including Go (reference level), high and low anticipation trials [Stop and No-Go trials were excluded]. Trial type was modeled as a 3-level factor with a priori non-orthogonal contrast coding, which compared high-anticipation vs Go trials and low-anticipation vs Go trials. Random intercepts were included for participants and for trial types. The effect of affective context on RTs was examined using the aforementioned approach. For Experiment 2, we implemented a similar analytic approach by fitting a linear mixed-effects model with main effects of trial type [high-anticipation vs Go, low-anticipation vs Go], session, age, and sex, and random intercepts for trial type and session were nested within the random effect of the participant. Session was included as either a main effect or interaction term, depending on L. Ratio test results. The final model was used for interpretation and affective context was included (again as either a main or interaction term based on L. Ratio test results).

To examine possible modulatory effects of affective load and further confirm the different nature of the inhibitory control components, accuracy across affective context images valences (positive, negative, neutral), with “neutral” set as reference was compared. When including the affective context term in the analyses, we tested via likelihood ratio test whether setting it as an interaction term would explain additional variance. Results from interaction effect models were only interpreted if this model explained additional variance when compared to the main effect only model via likelihood ratio test. For Experiment 2, accuracy was analyzed using a generalized linear mixed-effects model with a binomial distribution, including trial type (Go, No-Go), session, along with age and sex as covariates and a random intercept for participant. The inclusion of session as a fixed effect vs an interaction term was again determined via likelihood ratio test. The final model was retained for interpretation, and affective context was added to this model (either as a main or interaction effect, depending on L. Ratio test results).

Performance across sessions was compared using independent repeated measures ANOVAs with metric of interest measuring reactive and preemptive inhibition (SSRT, No-Go trial accuracy) as the dependent variable. Significant main effects were followed by pairwise paired-samples t-tests with Bonferroni correction for multiple comparisons (adjusted α = .017).

Changes in self-reported mental health symptoms between Session 1 and Session 3 were analyzed using Wilcoxon signed-rank tests, given the ordinal nature of Likert-scale responses. To control for multiple comparisons, False Discovery Rate (FDR) correction (Benjamini–Hochberg procedure, α = .05) was applied across the five primary PROMIS outcomes tested. Additionally, Spearman correlations were computed between change scores and age and sex to examine whether demographic characteristics were associated with symptom change. Changes in affect between Session 1 and Session 3 were examined using the Positive and Negative Affect Schedule (PANAS). For each of the 20 individual items, normality of change scores was assessed using D’Agostino’s K² test; parametric paired t-tests were applied when change scores were normally distributed (p > .05), and Wilcoxon signed-rank tests otherwise. Both Benjamini–Hochberg FDR and Bonferroni corrections were applied to account for multiple comparisons across the questionnaire items. Effect sizes were estimated using Cohen’s d. Spearman correlations were additionally computed between item-level change scores and age and sex. Analyses were also conducted at the subscale level, computing mean positive and negative affect scores (10 items each) per participant per session.

#### 2.3.5 Data and code availability

All data and code used in the present study are available from the corresponding author upon reasonable request.

## 3. Results

### 3.1 Experiment 1

#### 3.1.1 Participants

Among the initial 302 recruited participants, 241 completed at least 400 trials and responded to at least 70% of the Go trials. Following the filtering procedures described in section 2.3.1, 10 participants were excluded for not passing the attentional check and 63 for Stop trials accuracy response below 25% or above 75%, with 2 participants failing at both filtering criteria. One additional participant was excluded for instruction misinterpretation as they clicked the “A” key when the target stimulus was on the right side and vice versa, so that they had a Go trials accuracy rate of 4% and response rate of 82%. Following all exclusions, the final sample included 172 participants (age = 35.02 ± 11.42 years, males = 89).

#### 3.1.2 Preemptive and Proactive inhibition

To investigate preemptive inhibition, No-Go trials accuracy was analyzed with a model where Go trial accuracy was the baseline measure. There was a significant effect of trial type (β = 1.45, SE= 0.09, p < .001), indicating that accuracy was higher on No-Go as compared to Go trials, see Figure 2A, confirming the successful manipulation of preemptive inhibition. Age was associated with lower accuracy (β = -0.20, SE=0.07, p=0.005), indicating that younger participants tended to respond more accurately. Sex was not a significant predictor of accuracy (β = 0.24, SE=0.15, p = 0.11).

**Figure 2.**
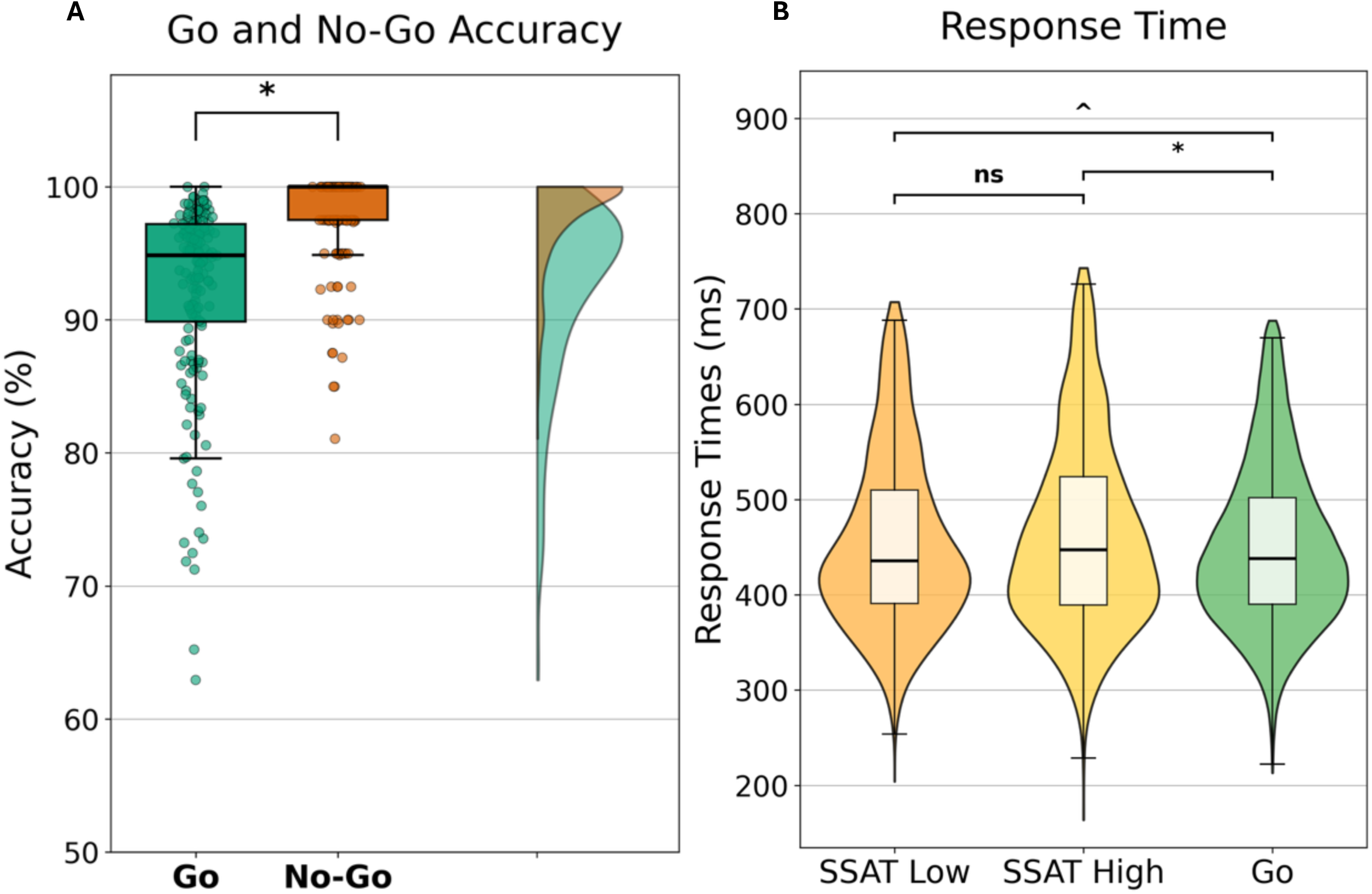
performance at preemptive and proactive trials. A: accuracy of Go vs No-Go trials. Percentage of correct responses of Go (left) and No-Go (right) trials showing diRerences between conditions. Boxplots show the median value (central black line), first and third quartiles (box edges), and range excluding outliers (whiskers), with individual participant data overlaid as points. On the right side, frequency distributions are depicted. **B: RTs at anticipation and Go trials.** Violin plots depicting RTs distributions across trial types (anticipation low, anticipation high and Go trials). Boxplots show the distribution median value (central line), first and third quartiles (box edges), and range excluding outliers (whiskers), with individual participant data overlaid as points.

When cued with an anticipation signal, participants generally slow their reaction to prepare to hold their response in case of a stop signal ^24,26^. In line with prior literature, RTs at high and low anticipation trials was analyzed to assess proactive inhibition, using Go trials RTs as the baseline condition. As predicted, there was a significant main effect of trial type, with slower responses for both high anticipation trials (β = 0.16, SE = 0.04, p < 0.001) and low anticipation trials (β = 0.07, SE = 0.04, p = 0.047) compared to Go trials, see Figure 2B. Age and sex were not significant predictors of RTs (p’s > 0. 202).

### 3.2 Experiment 2

#### 3.2. 1 Participants

A total of N=65 participants were recruited for Experiment 2. Exclusion criteria were applied on a session-by-session basis following the filtering procedures described in section 2.3.1. Of the participants who completed each session (Session 1: n=48, Session 2: n=46, Session 3: n=48), two participants were excluded from Session 2 and one from session 1 for not answering the attentional check question correctly. Additionally, when excluding those with Stop trials accuracy between 25 and 75%, for each session 15, 14 and 9 participant-session performances were excluded, respectively. Following all exclusions, our final sample comprised 39 unique participants (age = 40.39 ± 12.23 years old, females = 18, males= 15, unknown =6). Each session included 33, 31 and 38 participant-session files, respectively.

#### 3.2. 2 Preemptive and Proactive inhibition

For preemptive inhibition (Go vs No-Go accuracy), there was a significant main effect of both trial type and session on accuracy. Confirming the results from Experiment 1, accuracy was again higher on No-Go trials (β = 0.91, SE = 0.11, p < .001). It was also overall improved over sessions (β = 0.38, SE = 0.06, p < .001). A significant trial type × session interaction emerged (β = -1.17, SE = 0.25, p < .001), indicating that accuracy for No-Go trials diminished at Session 3, see Figure 3A. This pattern may reflect a shift in attention, with participants prioritizing more demanding trial types after task familiarization in session 1, leading to a relative decrease in No-Go accuracy. Older participants were less accurate (β = -0.39, SE = 0.22, p < .001). Sex did not significantly explain performance (all p’s>0.103).

**Figure 3.**
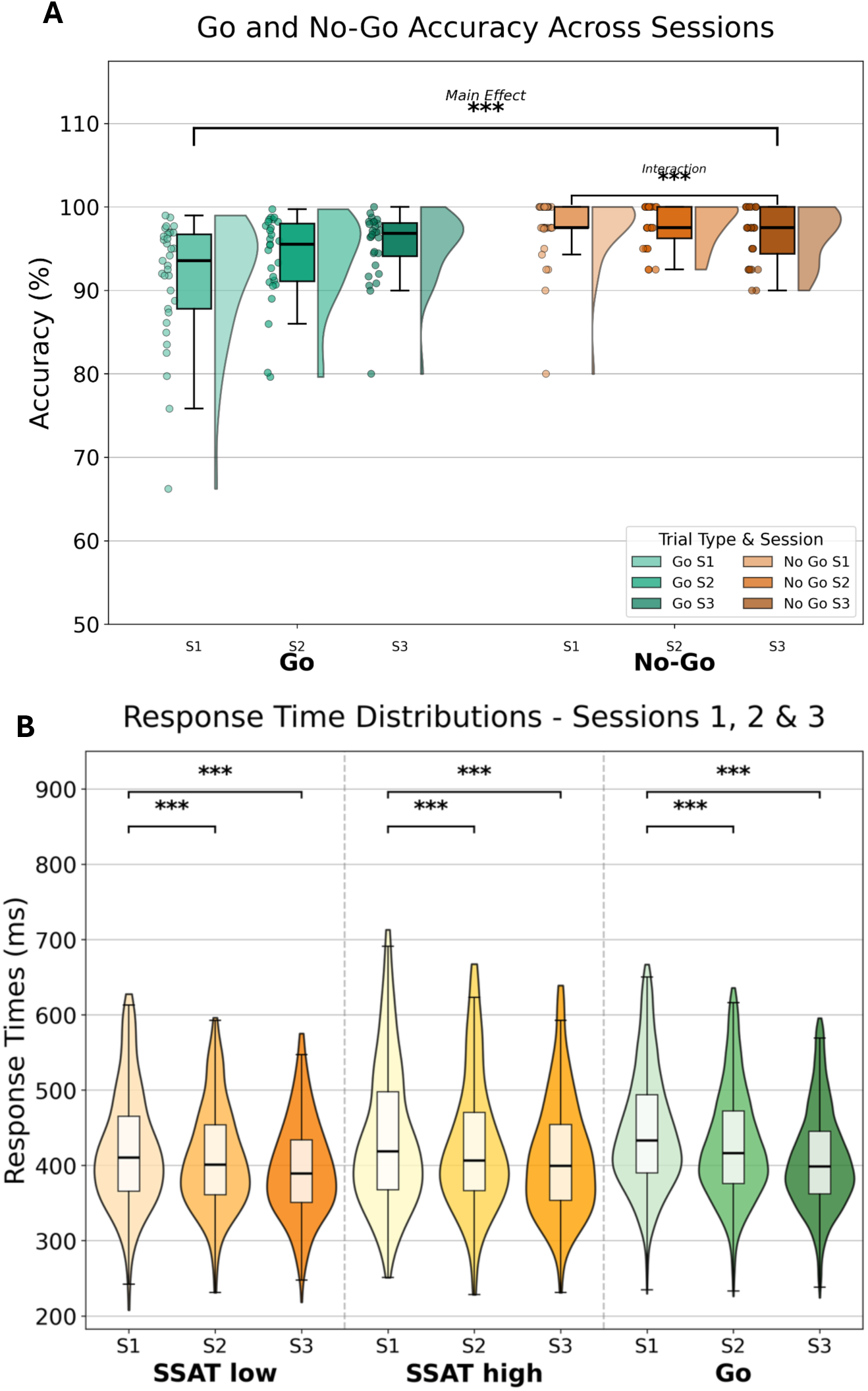
performance at preemptive and proactive trials across sessions. A: accuracy of Go vs No-Go trials. Percentage of correct responses of Go (left) and No-Go (right) trials showing diRerences between trial types and sessions. Boxplots show the median value (central black line), first and third quartiles (box edges), and range excluding outliers (whiskers), with individual participant data overlaid as points. On the right side of each boxplot, frequency distributions are depicted. **B: RTs at anticipation and Go trials across sessions.** Violin plots depicting RTs distributions across trial types (anticipation low, anticipation high and Go trials) and session. Boxplots show the distribution median value (central line), first and third quartiles (box edges), and range excluding outliers (whiskers), with individual participant data overlaid as points.

RTs at go, anticipation high and low trials were faster during sessions 2 and 3 compared to session 1 (session 2: β = -0.25, SE = 0.06, p < .001; session 3: β = -0.45, SE = 0.06, p < .001), indicating that participants became faster across sessions, see Figure 3B. Age and sex were not significant predictors of RTs (both p’s > .46). For anticipation trials, there were no effects of trial type, with high (β = -0.04, SE = 0.06, p = .461) and low (β = 0.005, SE = 0.05, p = .924) anticipation trials showing similar baseline RTs to the reference condition. This is at odds with our results in Experiment 1, possibly reflecting differences in statistical power associated with the smaller sample size of Experiment 2.

#### 3.2. 5 Task performance across sessions

Repeated measures ANOVAs revealed significant effects of session on inhibitory control measures. SSRT significantly decreased from Session 1 to 2 (F_(2,56)_ = 6.26, p = .004; post-hoc: t_(28)_ = 3.48, p = .002), indicating better reactive inhibition with practice (lower values indicate better inhibition), see Figure 4A. Additionally, No-Go trials accuracy decreased (F_(2,56)_ = 4.79, p = .012) from Sessions 1 to 3 (t_(28)_ = 2.74, p = .011) and from Session 2 to 3 (t_(28)_ = 2.64, p = .013), suggesting declining performance on these trials with practice, see Figure 4B. Overall, these results indicate that practice improved reactive inhibition, as reflected in faster SSRTs, but did not benefit preemptive inhibition, as shown by the decline in No-Go accuracy across sessions.

**Figure 4.**
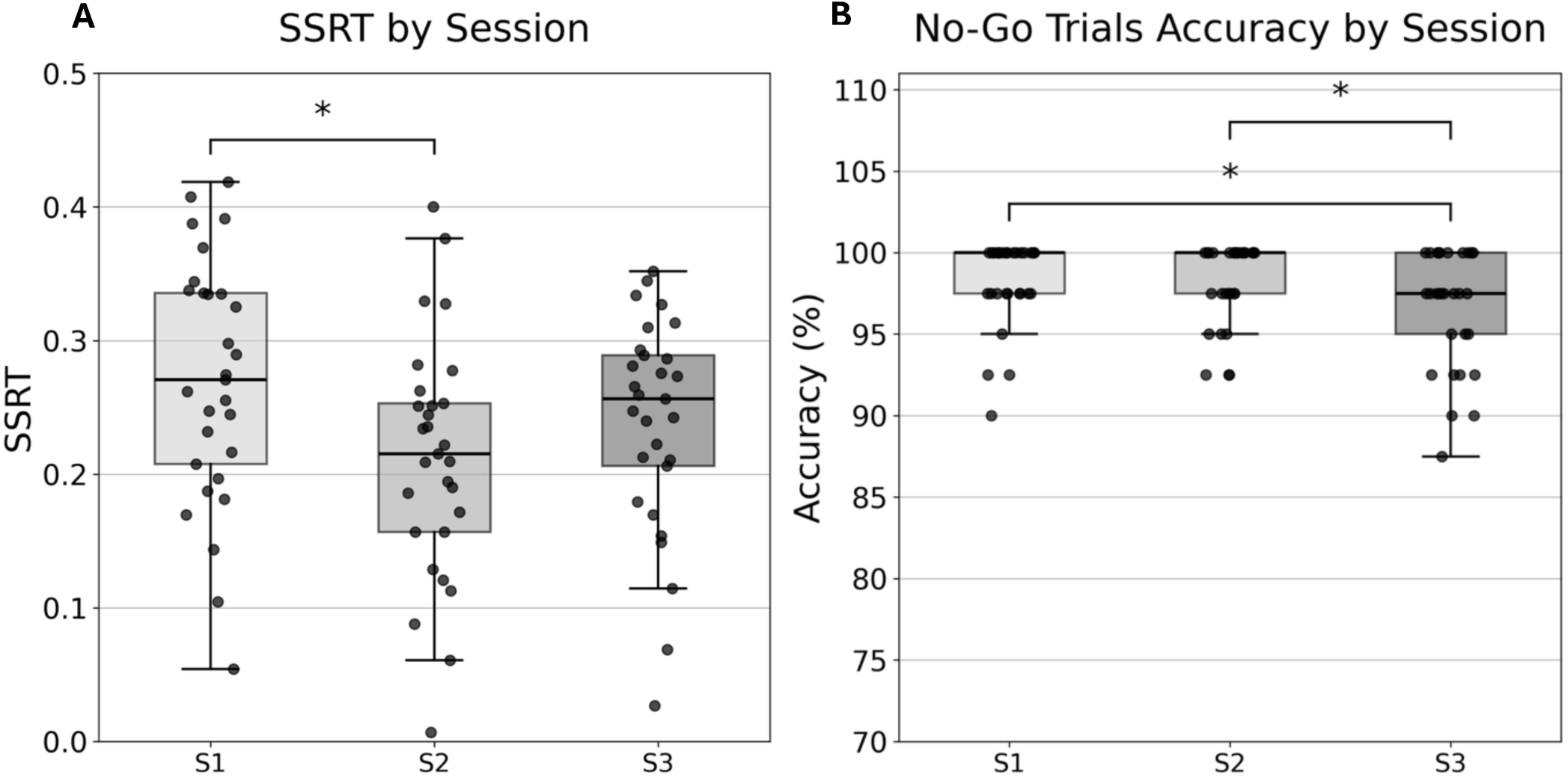
Task performance across sessions. **A:** SSRT value across sessions, lower values indicated greater reactive inhibition control. **B:** No-Go accuracy percentage across sessions. All boxplots show the median value (central line), first and third quartiles (box edges), and range excluding outliers (whiskers), with individual participant data overlaid as grey points. All boxplots show the median value (central line), first and third quartiles (box edges), and range excluding outliers (whiskers), with individual participant data overlaid as black points.

#### 3.2. 5 Symptoms improvement with training

To examine whether participants’ mental health symptoms changed over the course of the study, Wilcoxon signed-rank tests were conducted comparing selected PROMIS items investigating anxiety and temper control between Sessions 1 and 3 (n = 32 participants with complete data at both timepoints). Results revealed a decrease in self-reported anger ("I felt angry") from Session 1 (M = 2.25, SD = 0.76) to Session 3 (M = 1.84, SD = 0.77), W = 25.5, p = .007, q = .037, d = -0.51, see Figure 5A. No other PROMIS item showed significant changes (all p-values > .25). Additionally, to examine whether participants’ affective state changed over the course of the study, Wilcoxon signed-rank tests were conducted comparing all 20 PANAS items between Sessions 1 and 3 (n = 32 participants with complete data at both timepoints). One item reached the uncorrected significance threshold: self-reported enthusiasm ("I feel enthusiastic") showed a small increase from Session 1 (M = 2.41, SD = 1.14) to Session 3 (M = 2.66, SD = 1.29), W = 9.00, p = .046, d = 0.38. However, this effect did not survive correction for multiple comparisons (FDR-corrected q = .69). No other PANAS item showed significant changes (all uncorrected p-values > .09). At the subscale level, neither positive (M = 2.71 to M = 2.74, p = .658, d = 0.08) nor negative affect (M = 1.59 to M = 1.55, p = .390, d = -0.09) changed significantly across sessions.

**Figure 5.**
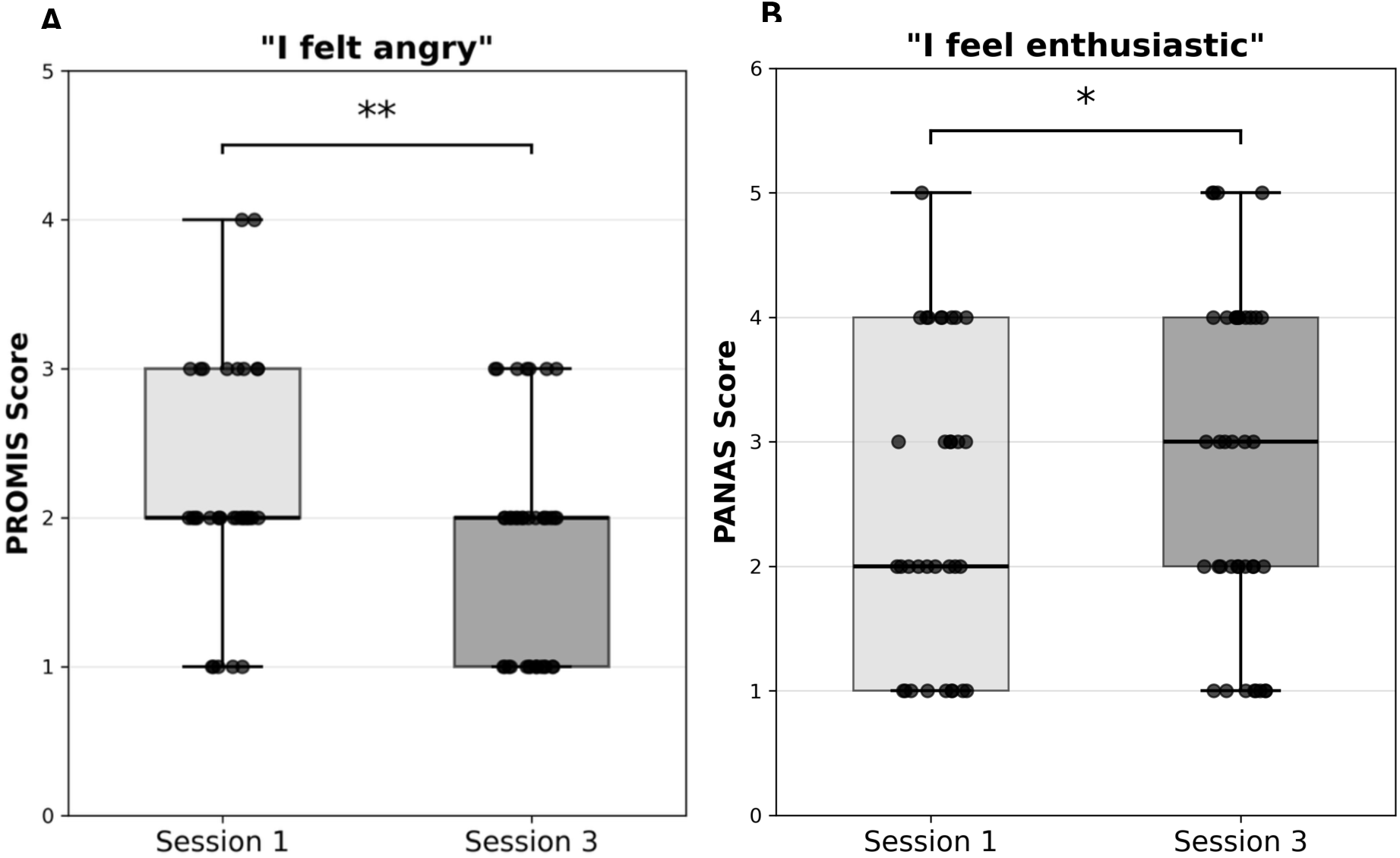
Mental health and aHective state before and after three GAMBIT sessions. **A**: self-reported anger values from PROMIS item. **B**: self-reported enthusiasm values from PANAS item. All boxplots show the median value (central line), first and third quartiles (box edges), and range excluding outliers (whiskers), and the individual participant data overlaid as black points.

The results described above were computed on the maximum data available, i.e. including all participant-session that passed the filtering step described in 2.3.1. In some cases, session 2 was not completed or did not pass the filtering criteria, the analyses were repeated only including participants who completed all three sessions and whose sessions were retained after filtering procedure (n=29). The same pattern of results was found in all models described above.

#### 3.2. 7 Usability

Participants completed a usability questionnaire at the end of Session 3, rating four aspects of the task on a 5-point scale. Overall, the task was perceived as moderately difficult (M = 3.87, SD = 1.36), easy to understand (M = 4.51, SD = 0.64), not particularly frustrating (M = 2.05, SD = 1.02), and participants indicated they would be willing to participate again (M = 4.13, SD = 1.42), see Figure 6. These ratings suggest that while the task was perceived as somewhat challenging, it was clear in its instructions, minimally frustrating, and generally well-received by participants.

**Figure 6.**
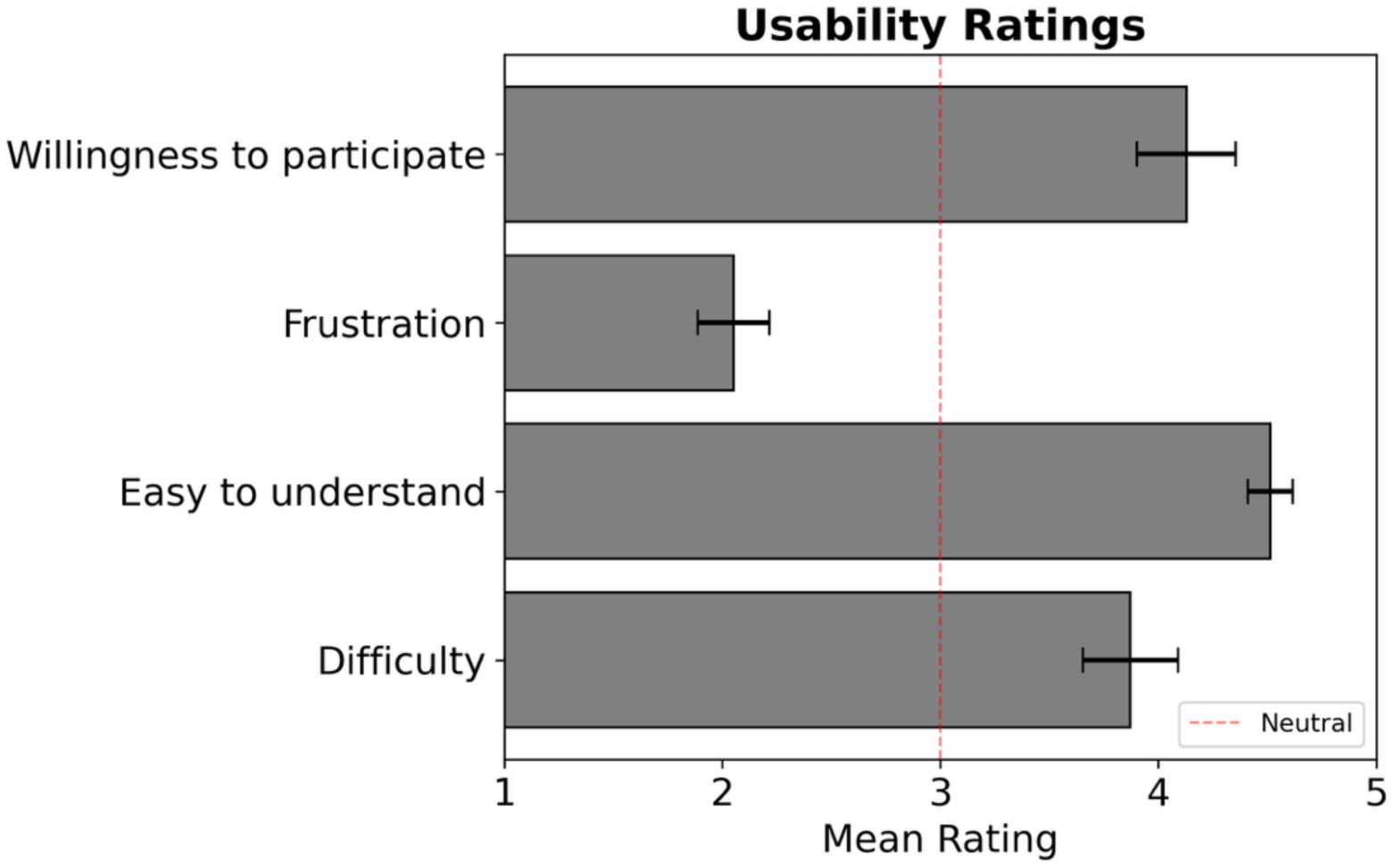
Barplot representing mean score per each question of the usability questionnaire. Each barplot represent the mean value for each usability question indicated on the left. Error bars represent error to the mean and red dotted line correspond to the median value 3.

## 4. Discussion

In the present study we describe a novel, fully online Inhibitory Control training paradigm designed to simultaneously engage preemptive, proactive, and reactive inhibition while incorporating emotional processing. Across two experiments, performance metrics targeting distinct inhibitory control processes revealed different patterns of results. Specifically, preemptive inhibition, measured via No-Go accuracy, was consistently different than reference Go trials in both experiments, indicating successful manipulation of this component. Proactive control, assessed through stop-signal anticipation trials, was reflected in slower reaction times compared with Go trials, particularly in the presence of affective stimuli, demonstrating that the task modulated proactive control and that emotional contexts can further interfere with this process. Reactive inhibition, measured via SSRT, improved across sessions, consistent with expected practice effect. Task performance and a self-reported measure of temper control improved with practice, supporting the feasibility and effectiveness of our online inhibitory control training paradigm. Usability ratings indicated that the task is challenging but manageable, enjoyable and clearly explained.

These findings support the view that inhibitory control comprises dissociable components, consistent with recent evidence from behavioral ^6^, brain imaging ^8^ and genetic studies ^7^. Our paradigm was designed to capture and train these components within a single task, and the differential patterns observed, together with their differential sensitivity to training, confirm their functional separability across preemptive, proactive, and reactive measures.

Contextual cues are often incorporated in paradigms to measure their influence on the main manipulation ^29,30^. Inhibitory control processes are often altered by affective elements ^21^ via mechanisms of perturbed attention and interference with the ability to inhibit responses ^31^. In our task, we observed this effect with higher RTs in trials containing positive and negative affective images, suggesting that emotional stimuli modulate inhibitory responding. Notably, the impact of affective elements did not change across sessions, suggesting that repeated practice improved inhibitory control without reducing the sensitivity to emotional interference. This discrepancy may reflect the sensitivity of the measurement tool rather than effect, consistent with prior work finding training effects in neuroimaging but not behavioral measures of emotional interference ^32^. Evidence from both clinical and subclinical samples indicates that affective interference with inhibitory control is exacerbated in individuals with depressive symptoms and with disorders marked by emotion dysregulation, including borderline personality disorder ^33^. In these populations, negative emotional contexts disproportionately disrupt inhibitory and attentional performance relative to healthy adults ^22,33^. Exploring these differences across clinical populations in future studies could provide valuable insight into the mechanism through which affective context interacts with inhibitory control.

Prior works suggests that repeated practice of Go/No-Go types of paradigms is associated with improvement in performance and with neuroplastic brain changes ^34,35^. In Experiment 2, our participants completed GAMBIT three times within one week, allowing us to assess relevant longitudinal behavioral effects. We observed improvements in accuracy (for Go, high and low anticipation trials), as well as faster RTs (for Go, high and low anticipation trials), indicating practice-related effects over time. Interestingly, results revealed a decline in No-Go accuracy, which may reflect fatigue in a trial type that requires less attention, as highlighted by the very high accuracy. The improvement in reactive, as well as proactive inhibition across sessions, suggests that the task effectively trains multiple aspects of inhibitory control. Importantly, in addition to behavioral improvements, participants reported reduced anger by Session 3. Although this finding requires further investigation to prove its causal relationship with the repeated use of GAMBIT, it aligns with prior evidence showing symptom improvement following inhibitory control training ^20^, suggesting that our task may also have beneficial effects on affective regulation.

This study was conducted fully online, a modality that has grown dramatically in both research ^36^ and clinical work ^37^ since the forced social isolation of the COVID-19 pandemic. Importantly, results from online experiments generally replicate those of in-person experiments, although often with smaller effects ^38^. Online testing offers several advantages, including the ability to reach participants from diverse backgrounds, including larger diversity of demographic characteristics. This is shown also with the recruitment platform we used here: Prolific ^39^. Conducting the study online was particularly appropriate for evaluating a training tool intended for remote administration, as it allowed us to confirm that the task functions reliably without direct supervision. Nevertheless, online administration also introduces limitations. Participants may have reduced attention or motivation, making it more difficult to ensure compliance in real time ^40^. Additionally, the absence of an in-person researcher prevents participants from asking for clarification, and the testing environment cannot be standardized across individuals. We addressed these challenges through extensive attention checks and filtering, which led to the exclusion of many performances (see sections 3.1.1 and 3.2.1 for Experiment 1 and 2 respectively) but ensured that the remaining data were of high quality. Despite these challenges, the ability to effectively administer interventions remotely remains a critical goal, particularly in psychiatry, where access to care is a major barrier ^41^. These limitations have driven the emergence of online interventions as useful tools to complement standard psychiatric treatments ^37^. Indeed, online training paradigms have increased in recent years to address accessibility barriers ^42^ and have demonstrated feasibility and efficacy ^17^ across several domains, offering a scalable, accessible, and flexible approach. Here, we developed a transdiagnostic inhibitory control training tool that can be administered fully online. Usability ratings suggest that the task was moderately challenging, clear, and minimally frustrating, with most participants willing to repeat it. These findings support its feasibility for remote administration and its potential integration into broader psychiatric care, providing a practical option for individuals with limited access to traditional treatments.

In summary, we show that GAMBIT, our novel, fully online inhibitory control training paradigm, effectively engages both proactive and reactive components of inhibitory control, it is associated with performance and temper control improvements, and is well-received by participants. The task captures and validates the multifaceted nature of inhibitory control, including its modulation by affective contextual factors. It demonstrates feasibility for remote administration and holds potential as a scalable, transdiagnostic tool for supporting cognitive and affective regulation. While further clinical testing is needed, these findings provide promising evidence that targeted inhibitory control training can enhance inhibitory performance and may contribute to symptom reduction.

## 5. Supplementary Material

### 5.1 Experiment 1: model comparison

The likelihood ratio test comparing model of Go / No-Go accuracy with and without interaction term with affective context indicated that the interaction model did not provide a significantly better fit to the data (χ²_(2)_ = 2.41, p = 0.300) when adding affective context. We thus retained the model investigating only main effects. When investigating RTs of high/ low anticipation trials vs Stop trials, model comparison via likelihood ratio test indicated that including affective context as interaction term improved model fit compared to a model with only fixed effects (L. Ratio _(4)_ = 35.53, p < .001). We thus retained the model with affective context interaction for subsequent analysis.

### 5.2 Experiment 1: stop trial accuracy

The model examining accuracy across stop trial types revealed higher accuracy on high (β = 1.14, SE = 0.053, p < 0.001) and low (β = 1.30, SE = 0.055, p < 0.001) anticipation compared to Stop trials. Negative (β = -0.25, SE = 0.053, p < 0.001) and positive (β = -0.16, SE = 0.054, p = 0.003) context were associated with lower accuracy compared to neutral. An interaction emerged between low anticipation trials and positive context (β = 0.41, SE = 0.137, p = 0.002), indicating that the negative effect of positive images on accuracy was attenuated in low anticipation trials compared to stop trials. No other interactions reached significance (ps > 0.08). Neither age (β = -0.007, SE = 0.034, p = 0.838) nor sex (β = -0.07, SE = 0.068, p = 0.283) significantly predicted accuracy.

**Figure S1.**
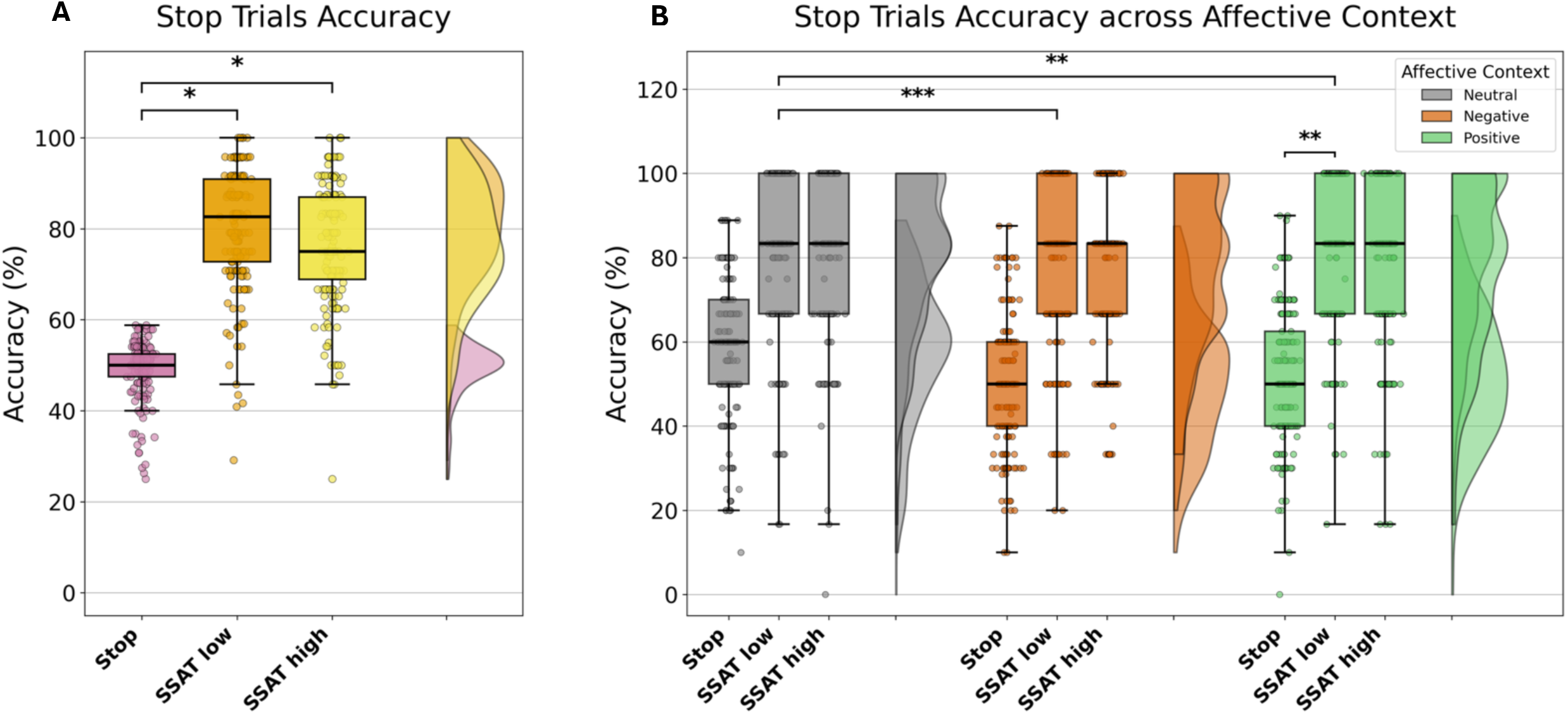
Accuracy at Stop trials across trial type and aHective context. **A:** Percentage of correct responses of Stop (left), SSAT low (middle) and SSAT high (right) trials showing diRerence between conditions. **B:** Percentage of correct responses across aRective context: neutral (left), negative (middle) and positive (right) across stop trial types (stop, SSAT low, and SSAT high). In both panels, boxplots show the median value (central line), first and third quartiles (box edges), and range excluding outliers (whiskers), with individual participant data overlaid as points. On the right-side frequency distribution are depicted.

### 5.3 Experiment 1: affective images

When testing the additional effect of affective load in the Go/No-Go model for preemptive inhibition, we found that affective context did not impact on accuracy, with no effect for negative (β = 0.07, SE = 0.042, p = 0.091) nor positive (β = -0.02, SE = 0.041, p = 0.610) images as compared to neutral, see Figure S2A. Age was associated with lower accuracy (β = -0.22, SE = 0.074, p = 0.004), indicating that younger participants tended to respond more accurately. Sex was not a significant predictor of accuracy (β = 0.25, SE = 0.149, p = 0.095).

When testing performance at the anticipation trials, we found that the positive affective context was associated with overall slower RTs as compared to neutral (β = 0.10, SE= 0.009, p < 0.001), while negative images showed no main effect (β = 0.008, SE = 0.009, p = 0.414). Additionally, interaction effects highlight that for high anticipation trials, both negative (β = 0.22, SE=0.072, p = 0.002) and positive (β = 0.33, SE= 0.07, p < 0.001) images were associated with slower RTs as compared to neutral. For low anticipation trials, the RTs were faster for positive images as compared to Go trials (β = -0.19, SE= 0.05, p < 0.001), while the negative image effect did not differ between low anticipation trials and Go trials (β = -0.08, SE= 0.03, p = 0.154), see Figure S2B. Age and sex were not significant predictors of RTs for both models (p’s > 0. 202). This effect suggests an additive/ larger cognitive load in trials including affective context images.

Notably, affective images influenced performance at anticipation trials but not No-Go trials, suggesting that preemptive and proactive inhibitory processes may be differentially sensitive to affective load, thereby confirming the multifaceted nature of inhibitory control as captured by our task. This pattern could also partially reflect differences in outcome metrics (RTs versus accuracy) used as the primary outcomes in the proactive and preemptive models.

**Figure S2.**
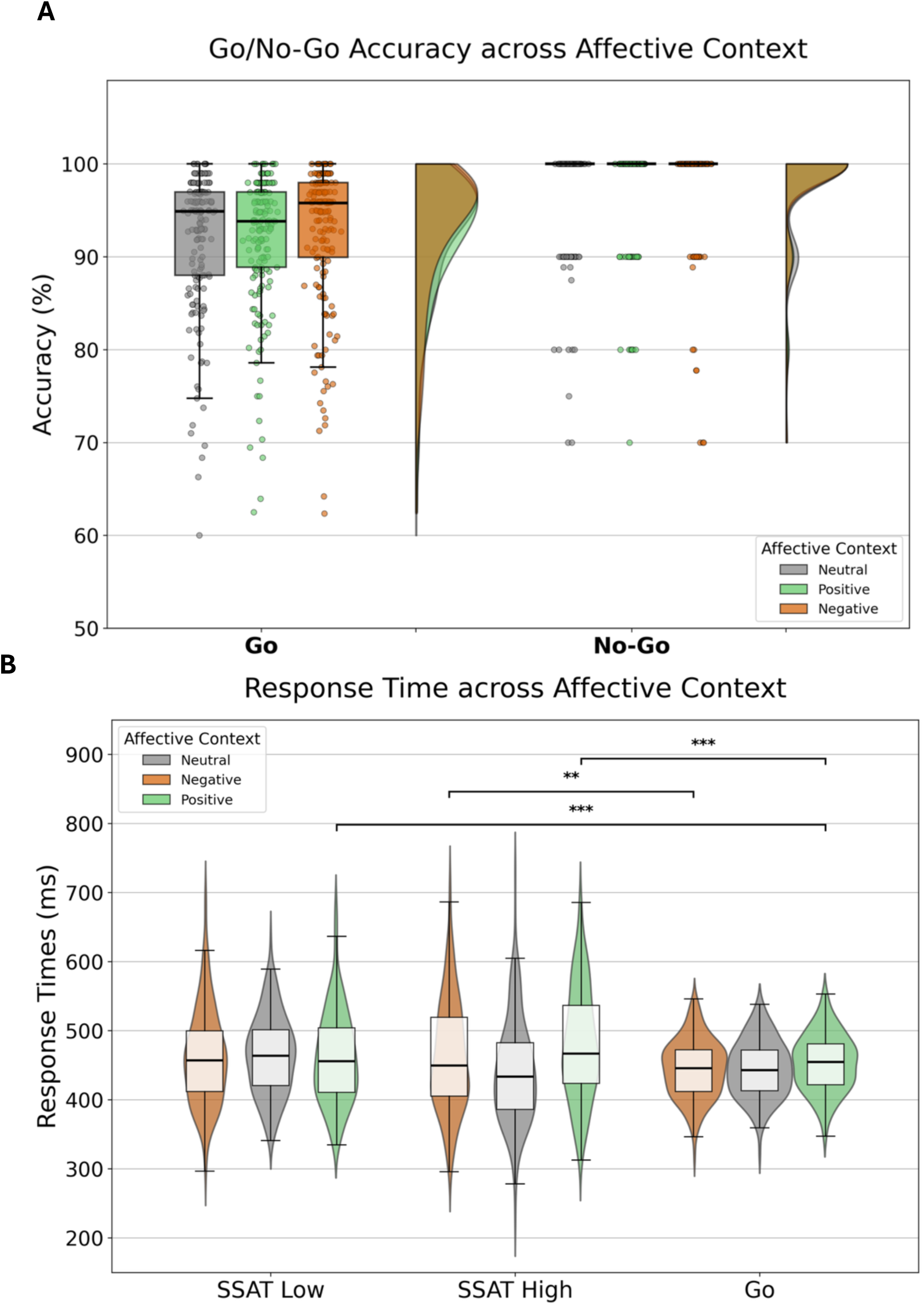
AHective images impact on No-Go and anticipation trials. **A:** Percentage of correct responses of Go (left) and No-Go (right) trials across neutral, positive and negative aRective contexts. In both panels, boxplots show the median value (central black line), first and third quartiles (box edges), and range excluding outliers (whiskers), with individual participant data overlaid as points. On the right side, frequency distributions are depicted. **B:** Percentage of correct responses of Go (left) and No-Go (right) trials across neutral, positive and negative aRective contexts. In both panels, boxplots show the median value (central line), first and third quartiles (box edges), and range excluding outliers (whiskers), with individual participant data overlaid as points. On the right-side frequency distribution are depicted.

### 5.4 Experiment 2: model comparison

When analyzing accuracy of Go/ No-Go trials, the inclusion of a trial type × session interaction term significantly improved model fit relative to the main effect only model (likelihood ratio χ²_(2)_ = 19.88, p < .001). We therefore report effects from the interaction model. Additionally, inclusion of the affective context interaction term in the model aforementioned improved fit relative to the main effect only model (χ²_(10)_ = 24.44, p = .007). As such, we report effects from this interaction effect model.

When analyzing RTs of high/low anticipation trials vs Stop trials, model comparison via likelihood improve model fit (likelihood ratio χ²_(4)_ = 7.43, p = .115). Similarly, the addition of an affective context-by-trial type interaction term did not improve model fit indices (likelihood ratio χ²_(8)_ = 12.20, p = .142), so results from the main effect only model were reported.

### 5.5 Experiment 2: stop trial accuracy

For the Stop, high and low anticipation trials models, the inclusion of affective context as interaction effect resulted in significantly improved fit indices as compared to the main effect only model (likelihood ratio χ²_(4)_ = 11.36, p = 0.023). Therefore, we retained the interaction model for interpretation.

**Figure S3.**
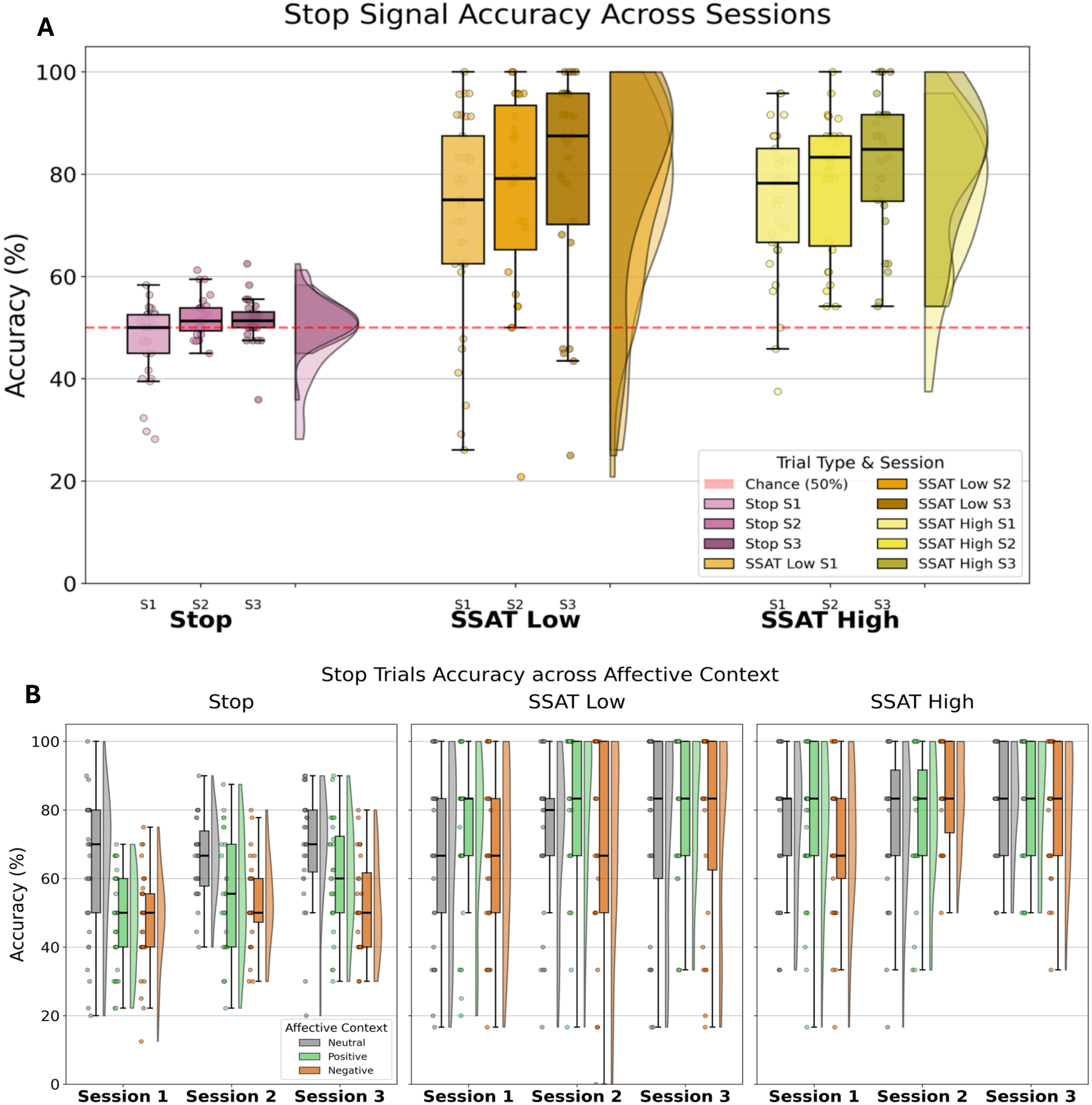
Accuracy at Stop trials across trial type, aHective context, and session. **A:** Percentage of correct responses of Stop (left), SSAT low (middle) and SSAT high (right) trials across sessions. **B:** Percentage of correct responses across aRective contexts and sessions: Stop (left), SSAT low (middle) and SSAT high (right) across aRective contexts and sessions. In both panels, boxplots show the median value (central line), first and third quartiles (box edges), and range excluding outliers

### 5.6 Experiment 2: affective images

When analyzing No-Go accuracy for preemptive inhibition, a significant interaction between negative affective context and Session 2 emerged (β = 0.35, SE = 0.15, p = .023), reflecting higher accuracy for negative images at Session 2. No other affective context × trial type interactions reached significance. No significant age or sex effect was (all p’s>0.103), see Figure S4A.

When analyzing anticipation trials RTs to investigate proactive inhibition, we found that affective images influenced performance. Compared to neutral images, negative ones were associated with slower RTs (β = 0.04, SE = 0.02, p = .039), and positive ones showed an even stronger slowing effect (β = 0.16, SE = 0.02, p < .001). We also observed an interaction effect between affective images and trial type, with negative affective images linked to slower RTs in both high (β = 0.27, SE = 0.11, p =.015) and low (β = 0.20, SE = 0.08, p = .017) anticipation trials. Positive affective images showed a similar pattern for high anticipation trials (β = 0.25, SE = 0.11, p = .023), whereas low anticipation trials showed faster RTs (β = –0.18, SE = 0.08, p = .025). Effects of affective images remained stable across sessions, with all image-by-session interaction terms showing no detectable changes (all p’s > .33), see Figure S4B.

**Figure S4.**
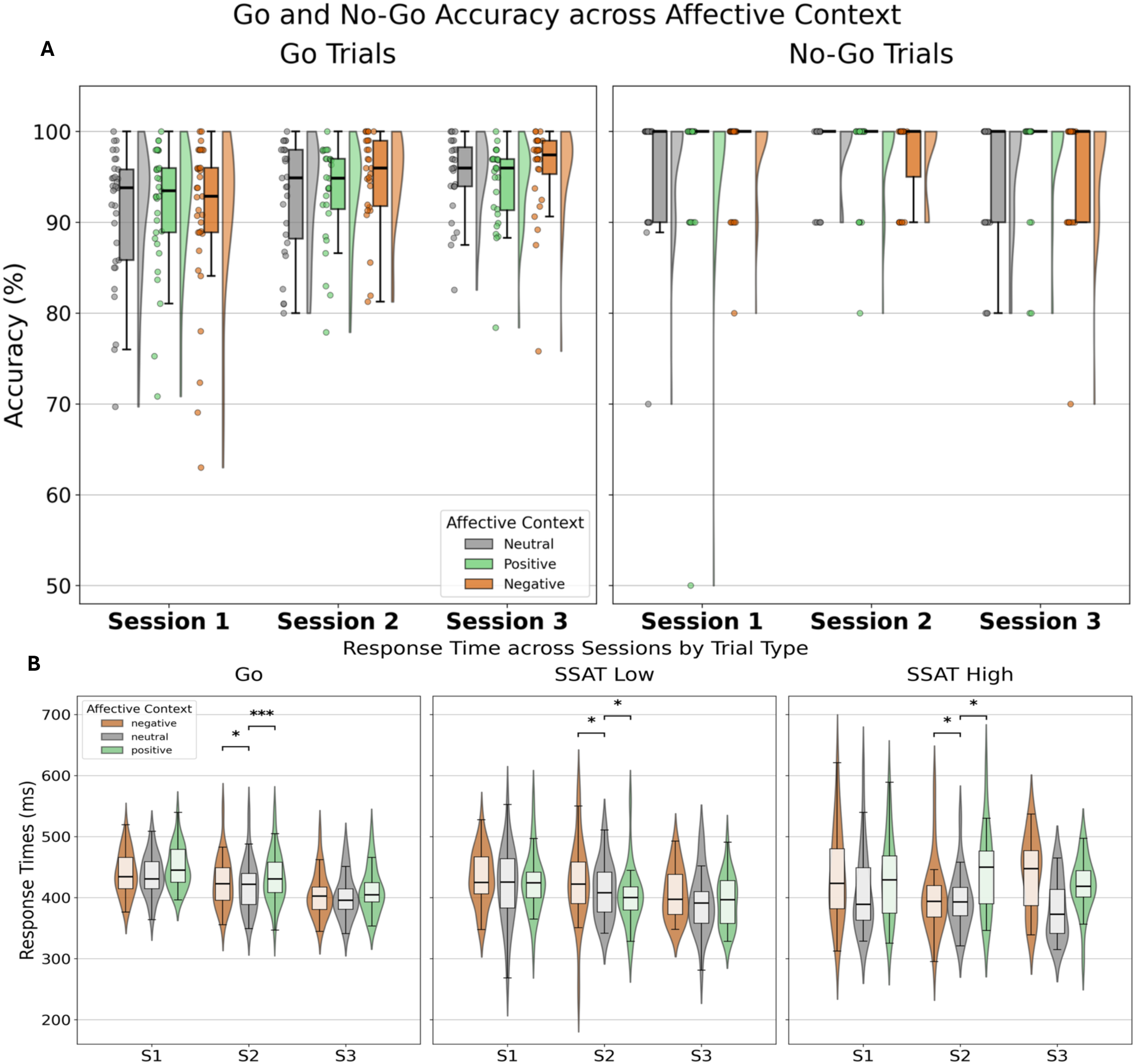
AHective images modulation of Preemptive and Proactive inhibition. A: Accuracy percentage across aRective context (Neutral, Positive, Negative) on Go trials (left panel) and No-Go trials (right panel). All boxplots show the median value (central line), first and third quartiles (box edges), and range excluding outliers (whiskers), with individual participant data overlaid as grey points. On the right side of each boxplot, frequency distributions are shown. **B:** Accuracy percentage across aRective context (Negative, Neutral, Positive) at Go trials (left panel), SSAT low (middle) and SSAT high (right). All boxplots show the median value (central line), first and third quartiles (box edges), and range excluding outliers (whiskers), with individual participant data overlaid as grey points.

## Author Contributions

Study conception and design: L.S.M., J.D.P.; Task development: L.S.M., G.D.; Data acquisition: G.D., S.K.; Statistical modeling: G.D., M.L.W.; Figures: G.D.; Drafting of the manuscript: G.D., S.K.; Critical revision of the manuscript: M.L.W., A.F., J.D.P., D.S.C., J.W.M., L.S.M.

## Acknowledgements

This work was supported by the Icahn School of Medicine at Mount Sinai, Mount Sinai Innovation Partners and in part through the generous support of the Ehrenkranz Center for Human Resilience at Mount Sinai, the Gottesman Foundation and Great Hill. This work was supported in part through the computational and data resources and staff expertise provided by Scientific Computing and Data at the Icahn School of Medicine at Mount Sinai and supported by the Clinical and Translational Science Awards (CTSA) grant UL1TR004419 from the National Center for Advancing Translational Sciences. We would like to thank the PsychoPy team, especially Dr. Rebecca Hirst.

